# Improved SARS-CoV-2 neutralization of Delta and Omicron variants of concern after fourth vaccination in hemodialysis patients

**DOI:** 10.1101/2022.06.22.22276787

**Authors:** Cho-Chin Cheng, Louise Platen, Catharina Christa, Myriam Tellenbach, Verena Kappler, Romina Bester, Bo-Hung Liao, Christopher Holzmann-Littig, Maia Werz, Emely Schönhals, Eva Platen, Peter Eggerer, Laëtitia Tréguer, Claudius Küchle, Christoph Schmaderer, Uwe Heemann, Lutz Renders, Ulrike Protzer, Matthias Christoph Braunisch

## Abstract

**Background:** Hemodialysis patients are exposed to a markedly increased risk when infected with SARS-CoV-2. To date it is unclear if hemodialysis patients benefit from a fourth vaccination.

**Methods:** A total of 142 hemodialysis patients (median age 72.6 years, 33.8% female) received four COVID-19 vaccinations between December 2020 and March 2022. RDB binding antibody titers were determined in a competitive surrogate neutralization assay. Vero-E6 cells were infected with SARS-CoV-2 variants of concern (VoC) Delta (B.1.617.2) or Omicron (B.1.1.529, sub lineage BA.1) in a biosafety level 3 laboratory to determine serum infection neutralization capacity before and after vaccination.

**Results:** After the fourth vaccination serum infection neutralization capacity significantly increased from a 50% inhibitory concentration (IC50, serum dilution factor 1:x) of 247.0 (46.3-1560.8) to 2560.0 (1174.0-2560.0) for the Delta VoC, and from 37.5 (20.0-198.8) to 668.5 (182.2-2560.0) for the Omicron VoC (each p<0.001). A significant increase of the neutralization capacity was even observed for patients who had high antibody titers after three vaccinations (p<0.001). Univariate regression analysis indicated immunosuppressive medication (p=0.001) and hepatitis B vaccination non-response (p=0.046), and multivariate analysis immunosuppressive medication as the only factor associated with a reduced effect against Delta (p<0.001). Ten patients with SARS-CoV-2 breakthrough infection before the fourth vaccination had by trend lower prior neutralization capacity for Omicron (p=0.051).

**Conclusions:** Our findings suggest that hemodialysis patients benefit from a fourth vaccination in particular in the light of the highly infectious SARS-CoV-2 Omicron variant. A routinely applied four-time vaccination seems to broaden immunity against variants and would be recommended in hemodialysis patients.

## Introduction

In hemodialysis patients, a SARS-CoV-2 infection is associated with a markedly increased morbidity and mortality in comparison to the general population with a mortality rate of more than 20% in hospitalized patients ^1-3^. In the last two years we have learned that double vaccination might not be enough to achieve adequate long-term immune protection in all hemodialysis patients and triple vaccination offers significantly better protection against COVID-19 in this patient group ^1, 4^. Even if an infection cannot always be prevented after booster vaccination, the course of the COVID-19 disease in general is milder depending on the number of vaccinations in hemodialysis patients ^5^. However, even in hemodialysis patients with an inadequate immune response after multiple vaccinations, morbidity remains significantly increased ^6^.

A third vaccination is associated with an increased virus neutralization capacity in the general population ^7, 8^.Therefore, to date, the third vaccination became part of the standard vaccination regimen, and meanwhile, a fourth vaccination is recommended in risk groups like hemodialysis patients as a so-called second booster ^9^. The usefulness of a third and now of a fourth vaccination is based on data from the general population and was obtained during the SARS-CoV-2 Delta wave. However, infections with the Omicron variant of concern (VoC) became predominant in 2022 ^10^. Omicron resulted in dramatically increased infection rates in the general population and in hemodialysis patients.

The question remains, whether the currently recommended vaccination regimen (four vaccinations, i.e. a second booster) in hemodialysis patients also offer effective protection towards VoC Omicron. It has not been clarified yet with certainty as to whether hemodialysis patients can further extend their immune response after a fourth vaccination. Interestingly, even if the immune response of a COVID-19 vaccination is less pronounced in hemodialysis patients compared to the normal population, there are indications that immune responses might be sustained in those hemodialysis patients who develop antibody responses ^11^.

The aim of this study was to investigate whether hemodialysis patients benefit from a fourth vaccination and if the immune response after the fourth vaccination has a comparable efficacy towards the VoCs Delta and Omicron. Furthermore, we examined the serum neutralization capacity of those hemodialysis patients who experienced a breakthrough infection after their third vaccination.

Here, we present the results of the live-virus infection neutralization of SARS-CoV-2 Delta and Omicron VoCs and antibody-mediated immunity before and after the fourth COVID-19 vaccination in a cohort of 142 hemodialysis patients.

## Material and Methods

### Study design

The COVIIMP study (German: “COVID-19-Impfansprechen immunsupprimierter Patient*innen”) is a prospective observational study examining the COVID-19 immunization success and the clinical course of COVID-19 in immunocompromised patients who received active or passive immunization against SARS-CoV-2 as recommended by the German health authorities. Patients were enrolled between April 1^st^, 2021 and March 20^th^, 2022. All patients are immunocompromised due to immunosuppressive medication after kidney transplantation, a rheumatologic disease demanding immunomodulatory therapy or End Stage Kidney Disease (ESKD) requiring maintenance hemodialysis.

The study is conducted at the university hospital Klinikum rechts der Isar of the Technical University of Munich in Munich and collaborating outpatient care centers. All participants provided written informed consent. The study, conforming to the ethical guidelines of the Helsinki Declaration, was approved by the Medical Ethics Committee of the Klinikum rechts der Isar of the Technical University of Munich (approval number 163/21 S-SR, March 19^th^, 2021) and registered at the Paul Ehrlich Institute (NIS592).

### Study population

Of 513 enrolled patients, a total of 142 patients requiring maintenance hemodialysis was selected. These patients received four COVID-19 vaccinations between December 29^th^, 2020 and March 20^th^, 2022 and underwent blood analysis before and after the fourth vaccination (**Figure 1A**). This subpopulation was recruited in four dialysis centers (Klinikum rechts der Isar, KfH Kidney Center Traunstein, Kidney Center Eifeldialyse, KfH Kidney Center München-Harlaching).

**Figure 1.**
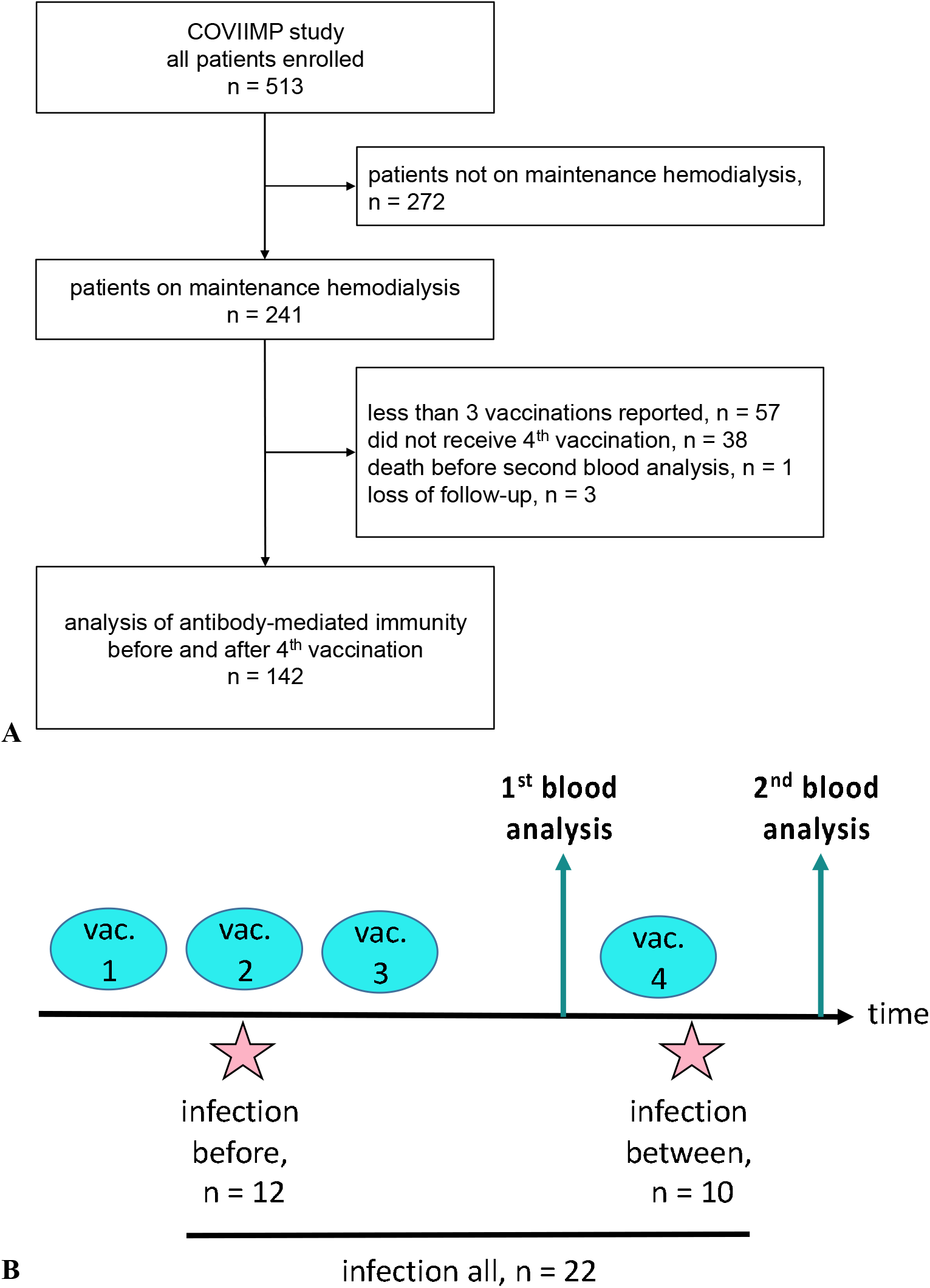
Flow chart of the COVIIMP study (A). Study design and observed SARS-CoV-2 infection cases (B). Abbreviations: vac., vaccination

Demographic data, medical history including current dialysis vintage, underlying kidney disease, history of transplantation and comorbidities as assessed by the Charlson Comorbidity Index (CCI) were collected. Immunosuppressive medication during the vaccination period was documented.

### Hepatitis B vaccination

Hepatitis B vaccination status was based on medical reports and, if available, serological laboratory data on anti-HBs antibodies. Patients were considered as non-responder if an anti-HB titer below 10 IU/l despite three hepatitis B vaccinations was documented or if their treating physicians classified them as a hepatitis B non-responder, according to local standards.

### SARS-CoV-2 infection

We identified participants as SARS-CoV-2 convalescent, if they had a prior positive SARS-CoV-2 PCR or at least one positive serological SARS-CoV-2 nucleocapsid-specific IgG measurement ^4, 12^. The clinical course and treatment were documented by structured interrogations.

### Sample collection

Blood was collected for analysis in median 2 (2.0 - 3.25) days before (analysis 1) and 26 (26.0-26.0) days after (analysis 2) the fourth vaccination.

### SARS-CoV-2 IgG assay

Antibodies in patients′ sera were detected using commercial surrogate paramagnetic particle chemiluminescence immunoassays (CLIA,Yhlo Biotechnology, Shenzhen, China) performed on the iFlash 1800 platform. Nucleocapsid-specific IgG antibodies (anti-N IgG) were determined using the 2019-nCoV IgG kit. The surrogate neutralization assay (NAb) was performed with the iFlash 2019-nCoV NAb kit based on the competition of serum antibodies with recombinant angiotensin-converting-enzyme 2 for binding the SARS-CoV-2 Wuhan strain receptor binding domain (RBD) and has been adapted for quantification to manufacturer’s instructions ^12, 13^. The cut-off level for seropositivity was set at 10 neutralizing units per milliliter (AU/ml) according to manufacturer’s instructions. Surrogate neutralization activity expressed as AU/ml can be adapted to WHO standard (AU/ml x 2.4 = BAU/ml [binding units / ml]). The maximum measurable value for NAb was 800 AU/ml, lower level of detection was 4 AU/ml. If values exceeded the upper limit of quantification a value of 801 AU/ml was used for statistical analysis. NAb high-response was defined as levels ≥700 AU/ml before the fourth vaccination. N-specific IgGs ≥10 AU/ml were qualitatively determined as reactive.

### SARS-CoV-2 infection-neutralization assay

Serum infection-neutralization capacity was analyzed as previously described ^8^. Briefly, SARS-CoV-2 isolates, which were kindly provided by Prof. Oliver Keppler’s group from the Institute of Virology and the Max von Pettenkofer Institute in Munich, were isolated from nasopharyngeal swabs of COVID-19 infected individuals. To obtain high titer of virus stock, Vero-E6 cells were infected with VoC Delta (B.1.617.2, GISAID EPI ISL: 2772700) or Omicron (B.1.1.529, sub lineage BA.1, GISAID EPI ISL: 7808190) and incubated in Dulbecco’s modified Eagle’s medium. After 2-3 days inoculation, cell culture medium was collected, centrifuged and the virus-containing supernatant was stored at −80°C. Prior to the neutralization experiments, viral titers were verified by plaque assay and strain identity was confirmed by next-generation sequencing. All measurements were performed using serum samples that were stored at −80°C and defrosted and stored at 4°C on the day before the analysis. Samples from all patients were analyzed in parallel. For quantification of the neutralization capacity, two-fold serial dilutions of the sera from 1:20 to 1:2560 were incubated with a predefined multiplicity of infection (MOI) of 0.03 (450 PFU/15,000 cells/well) of either of the VoCs for 1 hour at 37°C. The MOI was determined from an in-cell ELISA pre-test by which we observed viral signal saturation 24 hours after infection. After the 1-hour inoculation, the inoculum was transferred onto pre-seeded Vero E6 cells for another one-hour incubation at 37°C. The infection was terminated after one day and followed by an in-cell ELISA for the detection of SARS-CoV-2 N-protein. Cells were fixed with 4% paraformaldehyde and permeabilized by 0.5% saponin buffer. After blocking with 10% goat serum, cells were stained using anti-SARS-CoV-2-N primary (40143-T62, Sino Biological) and goat anti-rabbit IgG2a-HRP secondary antibody (EMD Millipore / #12-348), and eventually transformed into luminescence signal by adding substrate tetramethylbenzidine (TMB). To determine serum IC50 values, a nonlinear regression curve was applied and the dilution factor at which 50% inhibition was observed and was calculated using PRISM software (GraphPad). Patients were classified as low or non-responder if the IC50 value of the infection neutralization was ≤1:20.

### Statistical Analysis

Categorical variables are presented as frequencies and percentages. Continuous variables are expressed as mean ± standard deviation (SD) or median and interquartile range (IQR), as appropriate. Group differences were tested with the χ^2^ test or Fisher test. The independent samples *t*-test or Mann-Whitney-U test was used for continuous variables, as appropriate. Paired samples were examined with the Wilcoxon test and the McNemar test, as appropriate. Spearman correlation was used for correlation analysis.

Univariate and multivariate linear regression models were applied to identify possible predictors of the infection-neutralizing capacity of VoC Delta or Omicron (IC50) out of the following candidate variables: age, dialysis vintage, presence of immunosuppression, comorbidities, and hepatitis B vaccination non-response. Possible predictors were preselected prior to the statistical analysis. Logistic regression was used to examine the neutralization capacity towards an infection with SARS-CoV-2.

All tests were conducted two-sided and p < 0.05 was considered significant. Statistical analysis was performed using R version 4.0.2 (R Foundation for statistical Computing, Vienna, Austria).

## Results

Overall, 142 patients on maintenance hemodialysis were included (**Figure 1A**). Patients had a median age of 72.6 (61.5-80.6) years. 48 (33.8%) patients were female. The median dialysis vintage was 48.9 (21.3 - 83.7) months. At the time of the first, second, third and fourth vaccination, 124 (87.3%), 125 (88.0%), 136 (95.8%) and 142 (100%) were on maintenance hemodialysis, respectively. Further details of patient characteristics can be found in **Table 1** for all patients, and stratified by infection neutralization response against VoC Omicron.

**Table 1.**
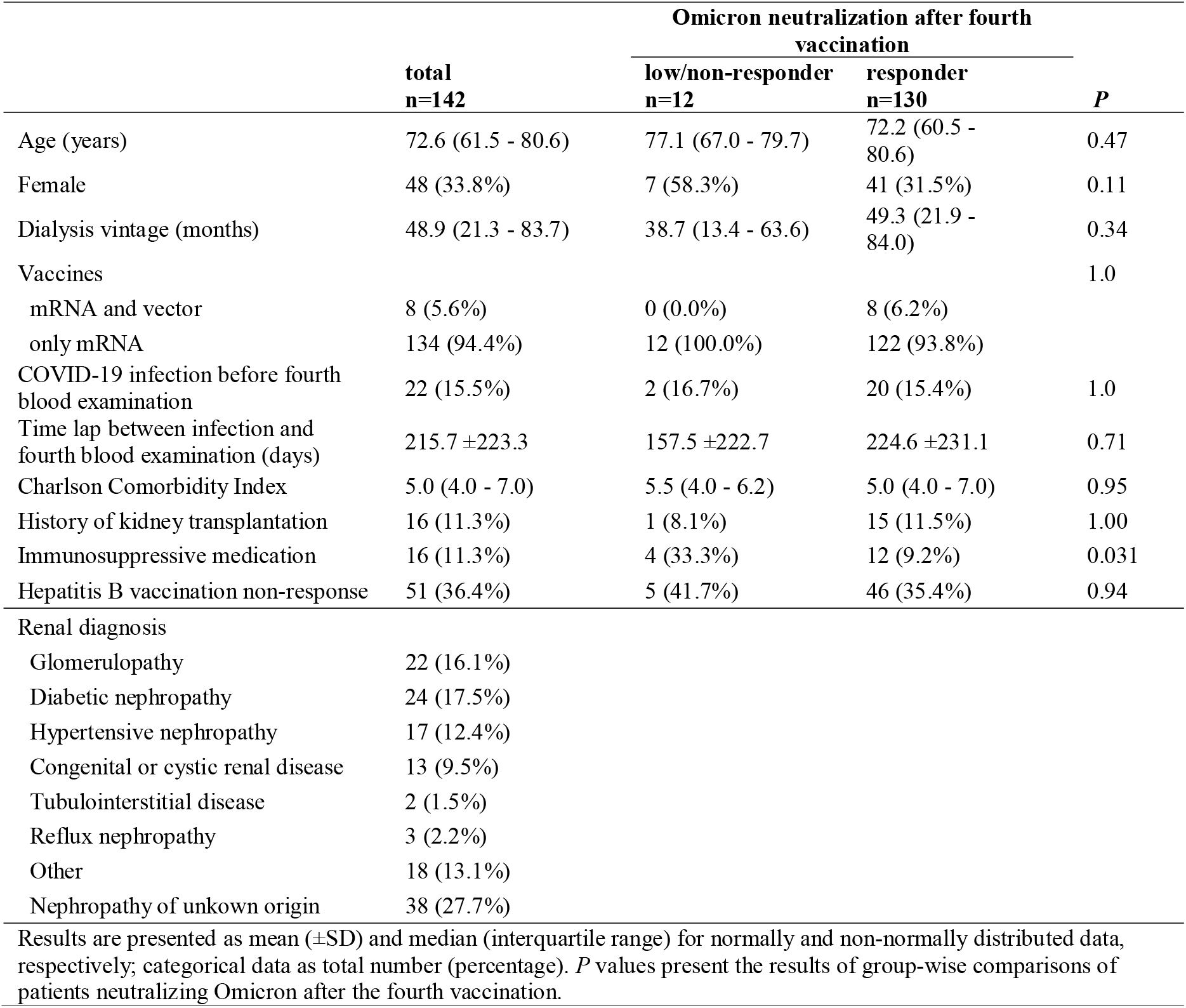
Patient characteristics.

### COVID-19 and vaccinations

All patients received four vaccinations. Eight and six patients received their first and second vaccination with AZD1222 (Vaxzevria^®^) by AstraZeneca, respectively. All other vaccinations were done with mRNA-based vaccines (BNT162b2 by BioNTech-Pfizer or mRNA-1273 by Moderna). Fifteen patients received two or more vaccinations with mRNA-1273 (Spikevax^®^, Moderna), the remaining patients received BNT162b2 (Comirnaty^®^, BioNTech-Pfizer). Median duration between the first and the fourth vaccination was 338.0 (333.0-342.0) days and between the third and the fourth vaccination was 126.0 (105.0-126.0) days, respectively. Median duration between the third vaccination and the first blood sampling was 4.1 (3.4-4.1) months.

A SARS-CoV-2 breakthrough infection indicated by SARS-CoV-2 nucleocapsid-specific IgG antibody positivity occurred in 22 (15.5%) individuals before the second blood sampling after the fourth vaccination (**Figure 1B**). In these patients, the average time between the SARS-CoV-2 infection and the second blood collection was 215.7 ±223.3 days. Of these, seven patients had no known history of SARS-CoV-2 infection, but were classified as convalescent due to positive anti-nucleocapsid IgG detection. Four (18.2%) of the 22 infected patients were treated with SARS-CoV-2 specific monoclonal antibodies. Ten (7.0%) patients had a SARS-CoV-2 infection between the two blood drawings before and after fourth vaccination. No patient reported recurrent SARS-CoV-2 infections (**Figure 1B**).

### Immunosuppression

Immunosuppressive medication was prescribed in 16 (11.3%) patients during the observation period. Reasons for immunosuppression were history of organ transplantation in eight, cancer treatment in four, underlying kidney disease in two and unknown causes in two other patients. Six patients received more than one immunosuppressive. Immunosuppressive agents were glucocorticoids in 14, tacrolimus in four, mycophenolate mofetil in three, and others in two patients (lenalidomide, rituximab and reduced dose CHOP).

### Impact of four vaccinations on neutralization capacity and NAbs

After the fourth vaccination significantly increased serum neutralization capacities were found for both VoCs, Delta and Omicron. Infection neutralization capacity for Delta increased from a median IC50 (serum dilution factor, 1:x) of 247.0 (46.3-1560.8) to 2560.0 (1174.0-2560.0), and for Omicron from 37.5 (20.0-198.8) to 668.5 (182.2-2560.0) (each p<0.001) (**Figure 2 A, B**). NAb levels significantly increased from 721.0 (184.5-801.0) to 801.0 (801.0-801.0, p<0.001) (**Figure 2C**). Serum neutralization capacity after the fourth vaccination was significantly lower for Omicron compared to Delta (668.5 [182.2-2560.0] vs 2560.0 [1174.0-2560.0], p<0.001). Similar to the overall cohort, when analyzing only NAb high-responder, we found a significant increase for the neutralization capacity for both VoCs, Delta (1172.5 [382.8-2560.0] vs 2560.0 [2560.0-2560.0], p<0.001) and Omicron (170.5 [56.3-468.5] vs 2553.0 [640.2-2560.0], p<0.001).

**Figure 2.**
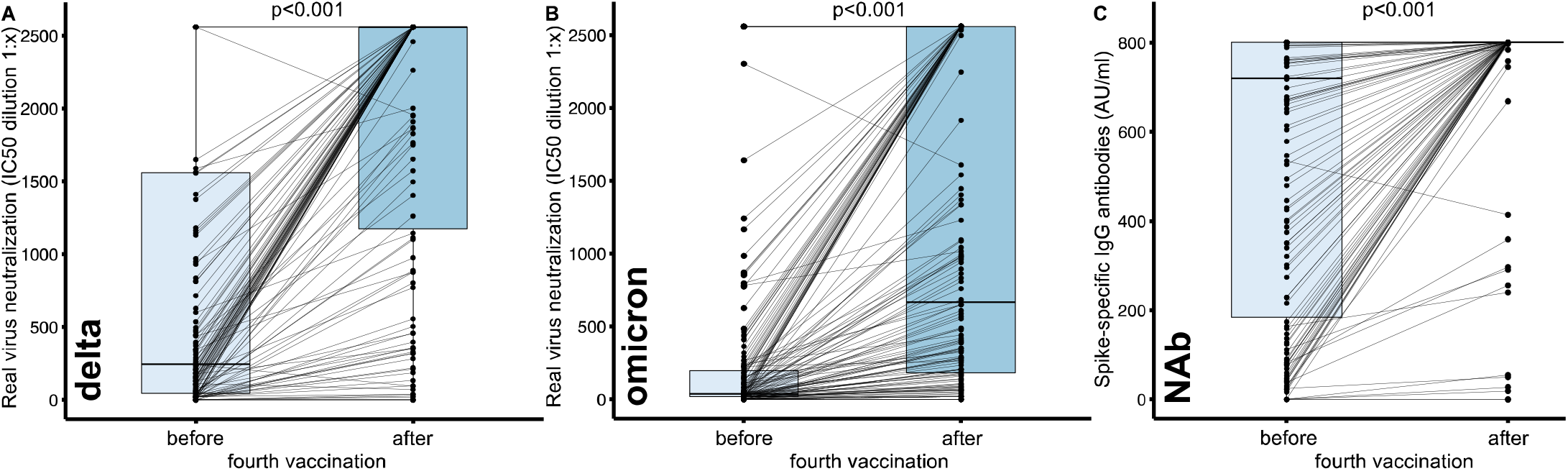
Changes of SARS-CoV-2 infection neutralization capacity before and after the fourth COVID-19 vaccination in hemodialysis patients. Real virus neutralization assay was performed using (**A**) the SARS-CoV-2 Delta (B.1.617.2) and (**B**) the Omicron (B.1.1.529, sub lineage BA.1) variant of concern upon serial dilution of hemodialysis patient sera before and after the fourth vaccination. Inhibitory concentration (IC50) dilution values are given. (**C**) Change of neutralizing antibody titers given in AU/ml in a surrogate neutralization assay. Dots indicate the measurement of an individual patient with lines connecting individual patient values before and after fourth vaccination. Boxes indicate median and interquartile range. Statistical analysis was performed using paired-samples Wilcoxon test, p values indicate statistical significance between groups.

Patients with a serum IC50 ≤20 were classified as low, those with no detectable neutralization as non-responder. Regarding both, Delta and Omicron infection neutralization capacity, significantly fewer patients (Delta: 30 vs 5; Omicron: 61 vs 12, each p<0.001) were low or non-responders after the fourth vaccination. The percentage of NAb responder was already very high before the fourth vaccination and did not further increase significantly (136 vs 139, p=0.13) (**Figure 3**).

**Figure 3.**
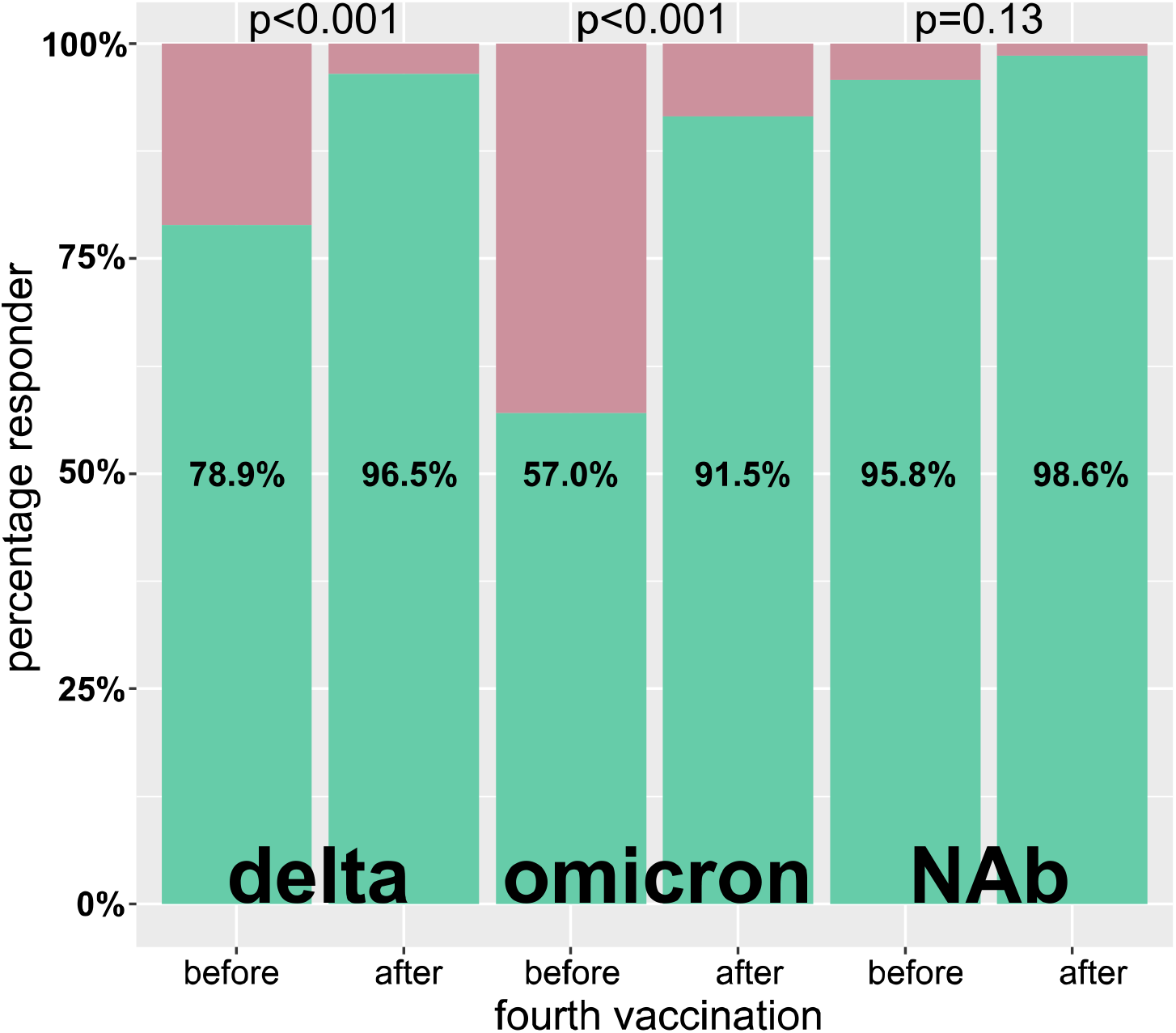
Percentage of responder before and after the fourth vaccination. A responder was defined by a Delta or Omicron IC50 virus infection neutralization of >1:20 as well as neutralizing antibodies (NAb) ≥10 AU/ml. Green and red indicate the percentages classified as responder and non-responder, respectively. Statistical analysis was done using McNemar test for paired samples.

After the fourth vaccination, infection neutralization of Delta and NAb titers were correlated highly significantly (p<0.0001) but moderately (rho=0.50) positive. Similarly, the correlation of the infection neutralization capacity of Omicron and NAb was highly significant (p<0.0001) and moderately (rho=0.44) positive.

Univariate regression analysis showed significantly reduced neutralization capacity for Delta after the fourth vaccination if immunosuppressive medication (p=0.001) or hepatitis B vaccination non-response (p=0.046) was present (**Table 2A, left column**). Multivariate analysis showed a reduced Delta neutralization capacity after the fourth vaccination if immunosuppressive medication (p<0.001) was taken and - by trend - if hepatitis B vaccination non-response was present (p=0.070) (**Table 2A, right column**). For Omicron infection neutralization, no such association was present in univariate or multivariate analyses (**Table 2B**). Univariate and multivariate analyses showed reduced NAbs after the fourth vaccination if immunosuppressive medication was prescribed (**Table 2C**).

**Table 2.**
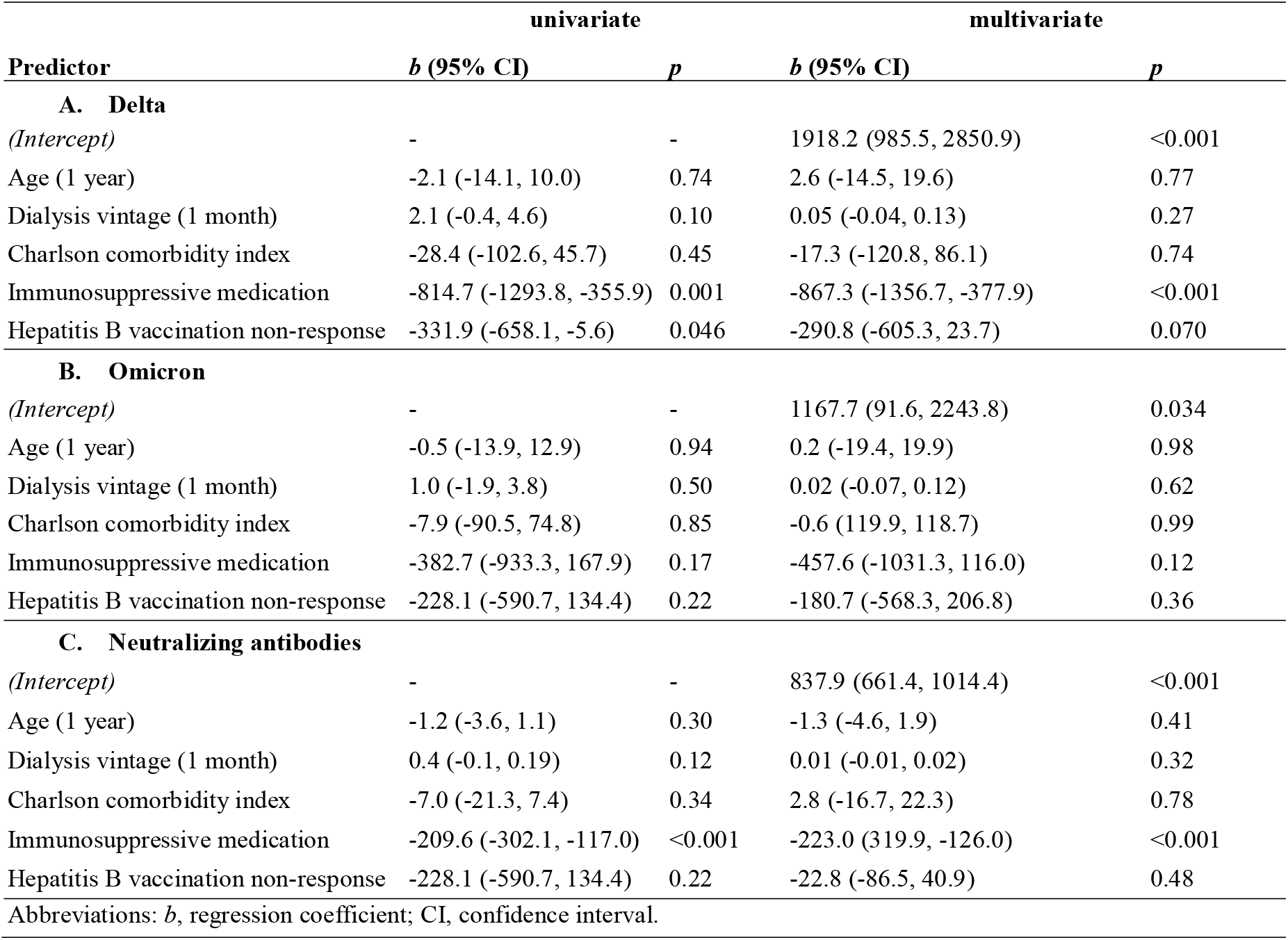
Univariate and multivariate regression models to identify predictors of Delta (**A**) and Omicron (**B**) neutralization capacity, respectively as well as neutralizing antibodies (**C**) after the fourth vaccination.

When comparing serum neutralizing capacities after the fourth vaccination between subgroups we saw significant differences in Delta infection neutralization if immunosuppression was prescribed (716.5 [176.2-2560.0] vs 2560.0 [1678.0-2560.0], p=0.002) (**Figure 4A**), and by trend for Omicron (193.5 [80.0-1481.8] vs 820.5 [214.3-2560.0], p=0.067) (**Figure 4B**). Patients with a history of SARS-CoV-2 infection had by trend a higher IC50 value for Delta (2560.0 [2560.0-2560.0] vs 2560.0 [955.2-2560.0], p=0.069) and significantly higher values for Omicron neutralization (1952.0 [893.2-2560.0] vs 489.0 [157.8-2560.0], p=0.013) (**Figure 4C, D**). If patients were classified as hepatitis B vaccine non-responder, they had significantly lower IC50 values for Delta neutralization (2460 [531.0-2560.0] vs 2560.0 [1765.0-2560.0], p=0.018) (**Figure 4E**), but not for Omicron neutralization (553.0 [103.5-1762.5] vs 760 [254.0-2560.0], p=0.18) (**Figure 4F**).

**Figure 4.**
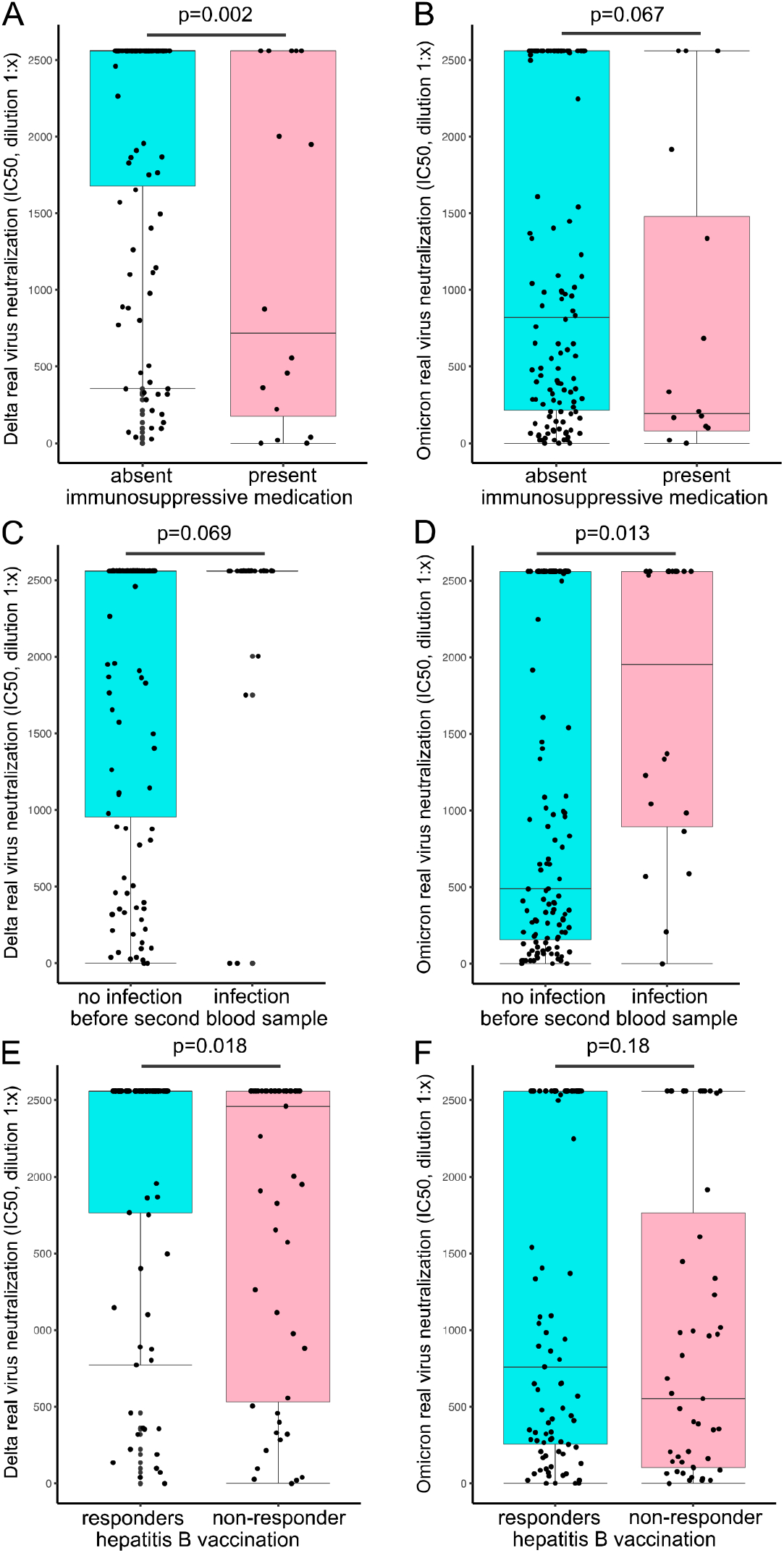
Influence of immunosuppressive medication, SARS-CoV-2 breakthrough infection and hepatitis B response status on COVID-19 vaccine responses. Serum real-virus neutralization capacity for Delta (left column) and Omicron (right column) was analyzed after the fourth vaccination in subgroups. Comparison of immunosuppressive drug treatment (**A, B**), prevalence of SARS-CoV-2 infection before the second blood sampling (**C, D**), and hepatitis B vaccination non-response (**E, F**) on serum neutralization capacity. Statistical analysis was performed using Mann-Whitney-U test, p values indicate statistical significance between groups.

### Impact of NAb and infection neutralization capacity on breakthrough infections

Finally, the ten patients with a SARS-CoV-2 infection before the fourth vaccination had by trend lower serum neutralization capacity for Omicron at the first blood sampling being almost significant (10.0 [0.0-26.8] vs 42.5 [20.0-217.5], p=0.051) (**Figure 5**). No difference was detected for serum neuralization capacity of Delta (189.5 [42.5-1167.0] vs 257.5 [50.8-1583.8], p=0.54). The VoC causing the SARS-CoV-2 infection was not determined. Omicron serum neutralization capacity after infection but before the fourth vaccination was not able to predict the COVID-19 breakthrough infection (p=0.29) when using logistic univariate regression.

**Figure 5.**
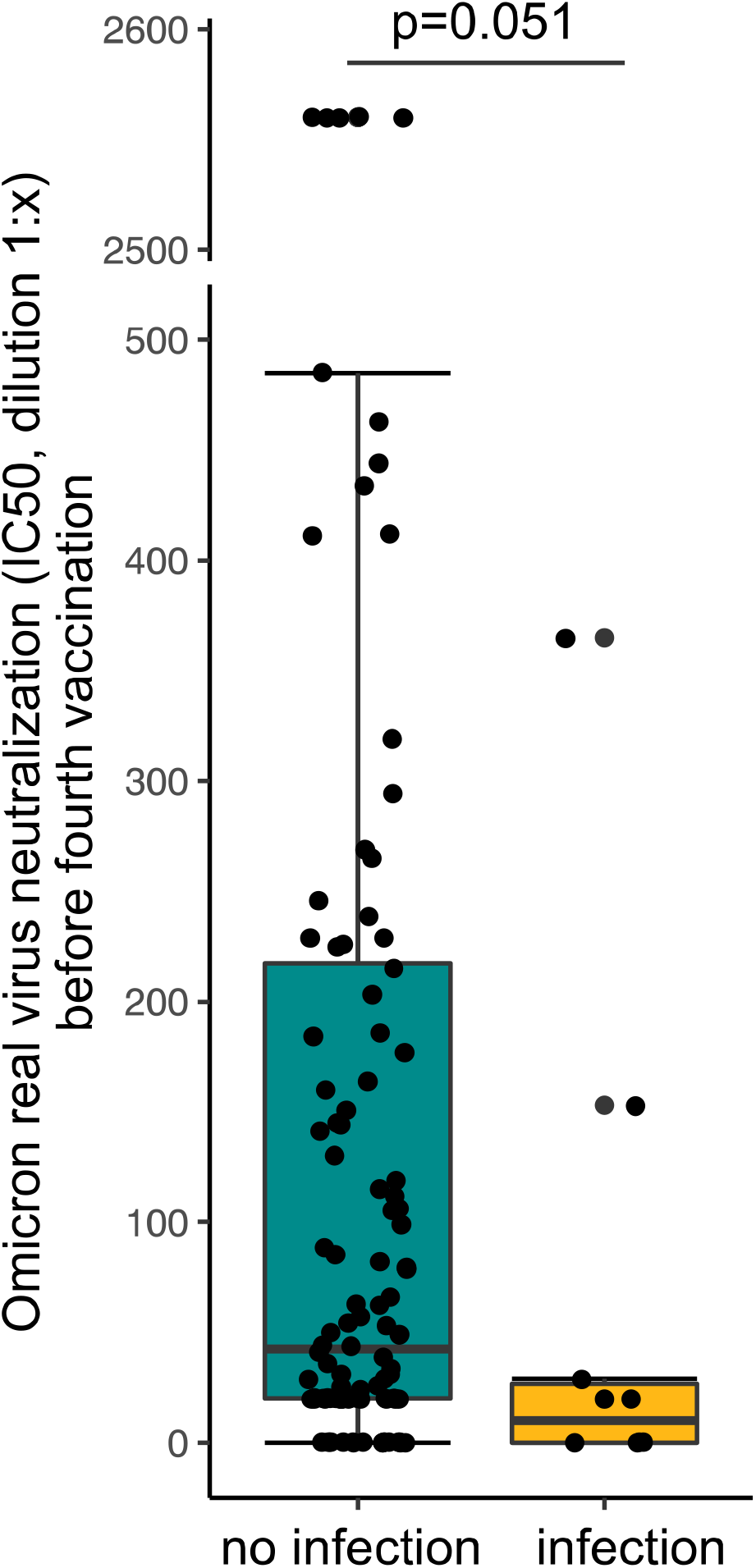
Serum neutralization capacity for Omicron variant of concern stratified by patients with SARS-CoV-2 breakthrough infections after the first blood sampling and before the fourth vaccination. Statistical analysis was performed using Mann-Whitney-U test, p value indicates statistical significance between groups. The y-axis is interrupted between 500 and 2500 for better visibility.

## Discussion

This prospective observational study demonstrates that hemodialysis patients benefit from a fourth COVID-19 vaccination. Serum infection neutralization capacity increased more than 10-fold for Delta and almost 18-fold for Omicron after a fourth vaccination indicating a better protection from infection with these highly infectious SARS-CoV-2 VoCs. The strength of our study is the examination of live-virus infection neutralization capacity of patients’ sera for the two most recent SARS-CoV-2 VoCs, Delta and Omicron. These two variants are also most distant from the original SARS-CoV-2 stain which was used to design the vaccines currently in use. Thus, the protective capacity against the new variants was hard to predict.

Our observation is of high importance since hemodialysis patients show reduced immunological responses to vaccination compared to healthy controls which may be explained in the context of uremia ^5, 14^. The hemodialysis patients in our study showed a significantly increased capacity to neutralize both SARS-CoV-2 VoCs, Delta and Omicron, after the fourth vaccination. This translates into significantly higher percentages of vaccine responders. Our results are consistent with previous reports of significantly increasing anti-spike antibody titers after the fourth vaccination in hemodialysis patients ^15, 16^, but add an important quality as these antibody titers were determined against the original vaccine stain of SARS-CoV-2 but not against the currently circulating variants. Furthermore, in line with a previous work with a pseudovirus assay we found a reduced neutralization capacity for VoC Omicron compared to Delta ^17^.

Patients with a breakthrough infection between the first and the second blood sampling had a lower neutralization capacity for Omicron only slightly missing significance. This was not seen for the Delta neutralization capacity. This might be at least partly explained by the fact, that the analysis was performed between February and March 2022 when the Omicron wave peaked in Germany. Hence, with over 99.3% the majority of COVID-19 cases were Omicron infections at that time ^10^. Logistic regression was not able to predict a SARS-CoV-2 breakthrough infection possibly due to the low infection rate after the first blood collection. In French hemodialysis patients a response towards wild type virus neutralization two weeks after the third vaccination was present in approximately 54% of patients ^5^. Another study in a British cohort found response rates of 97% and 72% for Delta and Omicron, respectively, in hemodialysis patients one month after the third BNT162b2 vaccination when applying an IC50 cut-off at 40 ^18^. We found response rates of 57% for Omicron and 79% for Delta four months after the third vaccination. Methodological differences in the neutralization assays ^5, 18^ as well as time interval differences associated with reduced immune responses to vaccination ^19^ might explain these variations.

In line with previous reports ^19-21^, we identified immunosuppressive agents as a predictor for lower neutralization capacity which were primarily prescribed to patients with a history of kidney transplantation. Patients on immunosuppressive medication had significantly lower neutralization capacity for Delta and, by trend, for Omicron. Other studies, however, did not identify immunosuppressive drugs as a predictor of neutralization capacity in hemodialysis patients ^5^. Discrepancies might be explained due to the specific immunosuppressive agents prescribed, as a previous study showed significantly reduced seroconversion rates in patients on anti-CD20 therapy regimes or mycophenolate mofetil, especially in the combination with glucocorticoids ^21^, substances which were also prescribed in our patients.

Interestingly, a positive hepatitis B vaccination response was by trend associated with an improved neutralization capacity. This was, however, only seen for the Delta VoC. It thus needs to be determined by further studies if hepatitis B vaccination response might serve as a surrogate for COVID-19 vaccination response or vice versa.

In clinical routine only NAb or anti-S antibody levels are readily and widely available. These, however, only detect the response against the original SARS-CoV-2 strains and not against the VoCs. Before the fourth vaccination, NAb were present in 96% of the study population and response rates did not further increase after the fourth vaccination. However, when looking at the absolute change of NAb titers, NAb increased significantly after the fourth vaccination. This increase was less pronounced than the increase in IC50 values in infection neutralization due to the limited range of the assay although the SARS-CoV-2 stain used for vaccination and in the NAb assay were identical. Although, NAb levels are regarded highly predictive of immune protection ^22^, this further demonstrates the limitation of the assays routinely available in the clinics.

We do not have outcome data of our cohort after the fourth vaccination with regard to infection prevention, but decreased COVID-19 incidence and severity in vaccinated hemodialysis patients has been observed by others ^5^. Thus, the presence of increasing NAb levels might still be a good indicator of vaccine response after the fourth vaccination and therefore useful in clinical routine. Nevertheless, further prospective studies have to evaluate how well a fourth vaccination protects hemodialysis patients from SARS-CoV-2 infection and COVID-19, respectively.

In a study by *Espi et al*., a third vaccination did not improve the immune response in patients that had already shown a high response after the second vaccination and was associated with more side effects ^5^. In our cohort, we did not record side effects but observed even in NAb high-responder a further significant increase of neutralization capacity and, more importantly, a very strong increase in infection-neutralization capacity of the two most prevalent SARS-CoV-2 VoCs. Differences worth mentioning to the work of *Espi et al*. might be the application of a third dose three months after the second dose. Whereas, the fourth vaccination was administered at least four months after the third dose in our cohort. Nevertheless, reports of increased side effects in high-responders may argue for an individual decision-making process depending on routinely available antibody levels.

Finally, some limitations have to be mentioned. We examined the neutralization capacity of the Omicron sub lineage BA.1. The question remains if these results are generalizable to other Omicron subvariants which are currently becoming predominant. Further studies have to show if improved neutralization capacity after the fourth vaccination is associated with COVID-19 incidence and severity.

## Conclusion

In conclusion, a fourth vaccination against SARS-CoV-2 significantly improves the antibody-mediated immune response in hemodialysis patients. A routinely applied four-time vaccination regimen therefore seems reasonable in hemodialysis patients. NAbs might be a good clinical surrogate of vaccination response. However, the presence of neutralization antibody titers above the upper limit of quantification should not hinder a fourth vaccination as this further improves and broadens live-virus infection neutralization. Further outcome data is necessary to evaluate the effect of a fourth vaccination towards SARS-CoV-2 breakthrough infection incidence and COVID-19 severity.

### Key learning points

*(3 bullet points per topic with max 50 words per point)*

### What is already known about this subject?

- Hemodialysis patients are a vulnerable patient group when infected with SARS-CoV-2, especially when also exposed to immunosuppressive medication.
- Four vaccinations increase neutralization antibody titers in hemodialysis patients.
- After a fourth vaccination, neutralization of the SARS-CoV-2 Omicron variant remained lower compared to the Delta variant in a pseudovirus assay.

### What this study adds?

- A fourth vaccination in hemodialysis patients largely (10- and 18-fold) improves the live-virus infection neutralization capacity for the most prevalent variants of concern, Delta and Omicron, respectively.
- Even in patients with high anti-S or neutralizing antibody titers binding the original virus, a parameter that is available in clinical routine, a further increase in neutralization capacity was demonstrated.
- Immunosuppressive medication was associated with reduced neutralization capacity for the Delta, but not the Omicron variant of concern.

### What impact this may have on practice

- A routinely applied four-time vaccination regimen seems reasonable in hemodialysis patients.
- A significantly improved neutralization of SARS-CoV-2 variants might warrant a fourth vaccination also in patients with high antibody titers in routine assays.

### Significance statement

Hemodialysis patients are a vulnerable patient group when infected with SARS-CoV-2 especially when also exposed to immunosuppressive medication. It is unclear if hemodialysis patients benefit from four vaccinations in terms of virus neutralization. We found a significantly improved infection neutralization capacity after the fourth vaccination in hemodialysis patients for both SARS-CoV-2 variants of concern, Delta and Omicron. Furthermore, also neutralization antibody high responder showed an improved and broadened virus neutralization capacity. Our findings suggest that hemodialysis patients benefit from a fourth vaccination against SARS-CoV-2, even when high neutralization antibody titers are already present. A routinely applied four-time vaccination regimen might be reasonable in hemodialysis patients.

## Data Availability

The datasets for this manuscript are not publicly available because written informed consent did not include wording on data sharing (German data protection laws). Reasonable requests to access the datasets should be directed to the corresponding author.

## Authors’ Contributions

CCC, LP, LR, UP, and MCB wrote the first draft of the manuscript. LP and MCB performed the statistical analysis. LP, MT, VK, CHL, MW, ES, EP, PE, LT, CK, CS contributed to blood sampling and data acquisition. CCC, CC, RB, BHL performed in vitro virus neutralization assays and measurement of neutralizing antibodies. Project supervision was done by LR, UH, UP, and MCB. Each author contributed important intellectual content during manuscript drafting or revision and accepts accountability for the overall work by ensuring that questions pertaining to the accuracy or integrity of any portion of the work are appropriately investigated and resolved.

## Acknowledgements

We would like to thank all patients for their participation in the study. We thank Prof. Oliver Keppler’s group from the Institute of Virology and the Max von Pettenkofer Institute, Ludwig-Maximilians University of Munich for kindly providing the SARS-CoV-2 isolates.

